# Improving the Detection of Mild Cognitive Impairment with FlowGAN: a Framework for ASL to FDG-PET Image Synthesis

**DOI:** 10.1101/2025.07.08.25331145

**Authors:** Chetan Vadali, Alfredo Lucas, T. Campbell Arnold, Sudipto Dolui, Sandhitsu Das, David A. Wolk, Kathryn A. Davis, Joel M. Stein, John A. Detre

## Abstract

**Background and Significance:** ^18^F-Fluorodeoxyglucose Positron Emission Tomography (FDG-PET) is commonly used to measure regional metabolism for diagnosis and monitoring of Alzheimer’s Disease (AD). However, FDG-PET is expensive and not widely available. Regional cerebral blood flow (CBF) is coupled to regional glucose metabolism and can be imaged noninvasively using Arterial Spin Labeling (ASL) perfusion MRI. Previously, we developed FlowGAN to synthesize FDG-PET images from ASL CBF and T1w MRI for lateralization of temporal lobe epilepsy (TLE). This study aims to extend FlowGAN to AD by examining the diagnostic potential of FlowGAN PET in mild cognitive impairment (MCI).

**Methods:** We included 47 MCI and 31 cognitively unimpaired (CU) individuals. FlowGAN, a generative adversarial network (GAN) for translating T1w MRI and ASL CBF maps into FDG-PET-like images, was trained using a 12-fold cross-validation scheme. We then evaluated the synthetic PET volumes by comparing them to true PET images both in appearance and for their classification performance in distinguishing MCI from CU via a random forest (RF) model, within regions of interest (ROIs).

**Results:** Synthetic FlowGAN PET volumes showed significant structural similarity to true PET volumes (SSIM = 0.958). Moreover, the performance of the best RF models for classification of MCI versus CU were comparable between the original PET and FlowGAN PET, both when considering all ROIs (PET AUC = 0.87, 95% CI: [0.79, 0.95]; FlowGAN AUC = 0.86, 95% CI: [0.77, 0.94]) and only a subset (PET AUC = 0.84, 95% CI: [0.77, 0.94]; FlowGAN AUC = 0.83, 95% CI: [0.74, 0.92]).

**Conclusions:** FlowGAN PET performs comparably to true PET in both appearance and classification. Since ASL can be acquired as part of a routine multimodal MRI protocol that is typically performed in patients with cognitive complaints, these findings may lead to improved access to diagnosis and treatment for AD.

## Introduction

Cognitive complaints are an increasingly frequent reason older adults seek medical attention, and this trend is expected to intensify with a growing aging population. These complaints can stem from a wide array of conditions, including Alzheimer’s Disease (AD), mild cognitive impairment (MCI), frontotemporal dementia (FTD), dementia with Lewy bodies (DLB), and various other neurological or psychiatric disorders. Accurately distinguishing among these potential diagnoses is essential, as management and prognosis vary considerably between AD and other dementias^1^. ^18^F-Fluorodeoxyglucose Positron Emission Tomography (FDG-PET), an imaging modality that measures glucose metabolism, is crucial for identifying characteristic patterns of hypometabolism associated with different dementias and detecting co-pathologies with AD^2–5^. However, the widespread use of FDG-PET is limited by its high cost and associated access challenges^6^.

Arterial Spin Labeled (ASL) perfusion MRI offers a non-invasive, MRI-based method of measuring cerebral blood flow (CBF) without radiotracers^7,8^. ASL sequences can be incorporated into standard MRI protocols commonly used for studies of neurodegenerative disease^8^, allowing CBF measurements to approximate glucose metabolism and support FDG-PET findings. In a recent meta-analysis, Haidar et al. reported variable comparability between ASL MRI and FDG-PET in dementia, likely reflecting the variable sensitivity and spatial resolution of ASL MRI technologies to date^9^. Further investigations have demonstrated similar performance in AD dementia diagnosis between FDG-PET and ASL, but stronger diagnostic potential with FDG-PET for amnestic MCI patients^10,11^. Therefore, FDG-PET is still the clinical modality of choice^9,11–17^.

Both FDG-PET and ASL MRI provide maps that reflect a measure of brain function (glucose metabolism or CBF, respectively) superimposed upon the underlying brain structure. Because ASL MRI is almost universally acquired as part of a multimodal MRI protocol that includes high sensitivity and high-resolution T1-weighted structural imaging, an opportunity exists to train deep learning methods to combine these modalities to generate PET-like images. Hussein et al. utilized convolutional encoder-decoder networks to synthesize ^15^O-water PET from multi-contrast MRI^18^, Chen et al. leveraged diffusion models for synthesis of FDG-PET from T1-weighted (T1w) MRI^19^, and Ouyang et al. applied convolutional neural networks to generate FDG-PET-like images from multi-contrast MRI, including raw ASL inputs^20^. In our prior work, we introduced FlowGAN, a deep learning framework designed to synthesize FDG-PET from paired T1w MRI and ASL CBF maps for temporal lobe epilepsy (TLE) patients^16^, which demonstrated strong results in capturing hypometabolism patterns critical for TLE localization. This approach—built on our LowGAN framework for low-to-high field MRI translation^21^—uses paired data to enhance the ability of the network to detect patient-specific pathology. Moreover, the use of both T1w MRI and ASL CBF—incorporating both structural and perfusion data—enables FlowGAN to better capture regional metabolic patterns critical for clinical interpretation.

In this study, we extended the application of FlowGAN to a cohort of patients with MCI. First, we assessed whether FlowGAN could generalize effectively to this novel patient cohort by generating synthetic FDG-PET images from MR-based structural and perfusion imaging data. Second, we evaluated whether these synthetic FDG-PET images exhibited hypometabolic patterns consistent with those observed in real FDG-PET images of MCI patients. Finally, we investigated the potential of synthetic FDG-PET images to accurately differentiate MCI patients from cognitively unimpaired participants based on regional patterns of flow/metabolism. We found that FlowGAN-generated synthetic FDG-PET images resembled true FDG-PET, recapitulated characteristic patterns of hypometabolism in MCI patients, and classified patients as MCI or CU based on imaging with similar precision to true FDG-PET, validating the robustness of this approach.

## Materials and Methods

### Study Design and Cohort Selection

This study used the dataset from Dolui et al.^14^, analyzing MRI and FDG-PET/CT data from 47 patients with mild cognitive impairment (MCI) and 31 cognitively unimpaired adults in the Penn Memory Center (PMC) cohort. The MCI diagnosis followed the National Institute on Aging – Alzheimer’s Association (NIAAA) criteria, based on consensus from specialists in neurology, psychiatry, radiology, and neuropsychology. Participants were aged 50-85, fluent in English, and had at least seven years of education. Exclusion criteria included a history of clinical stroke, traumatic brain injury, or other conditions impacting cognitive function. The study adhered to the Declaration of Helsinki and Institutional Review Board standards, with all participants providing written informed consent.

### Imaging Data Acquisition

#### FDG-PET Imaging

FDG-PET/CT data for 31 CU and 47 MCI patients were acquired using a Philips Gemini TF PET/CT scanner on the same day as their MRI scans. Participants fasted for at least 4 hours prior, with blood glucose levels verified to be under 180 mg/dL before receiving a 5.0 ± 0.5 mCi 18F-FDG injection intravenously while resting with their eyes open. A 30-minute 3D emission scan was performed 30 minutes post-injection, comprising six 5-minute frames (256 mm FOV, 128×128 matrix, 2×2×2 mm³ voxel size). A longer acquisition time compensated for the reduced tracer dose. Images were reconstructed using a line-of-response maximum likelihood algorithm with CT attenuation correction.

#### MR Imaging

The MRI data was acquired using a 3-Tesla Siemens Trio MRI scanner as part of a multimodal protocol. Arterial Spin Labeling (ASL) data was collected with pseudo-continuous labeling, using a 1.52-second labeling duration, a 1.5-second post-labeling delay, and a labeling plane offset of 9 cm. The acquisition included 45 label-control pairs with a 2D echo planar imaging (EPI) readout, TR/TE of 4 seconds/18 milliseconds, matrix size of 64×64, 3.4×3.4 mm² in-plane resolution, and 6 mm slice thickness with 20% spacing. Additionally, T1-weighted images were acquired using a 3D MPRAGE sequence (TR/TE/TI: 1.9s/2.89ms/900ms) at 1×1×1 mm³ voxel size across 176 slices.

#### Image Preprocessing

The ASL data underwent preprocessing with ASLprep^22^ for motion correction, estimation of CBF, and subsequent registration to the T1w MRI. Two sets of CBF maps were then created: one set was smoothed with a Gaussian kernel with σ=1, and the other set with σ=3 (**Figure 1A**). The FDG-PET data were registered and resliced to T1w volumes using ANTs affine registration^23,24^.

**Figure 1.**
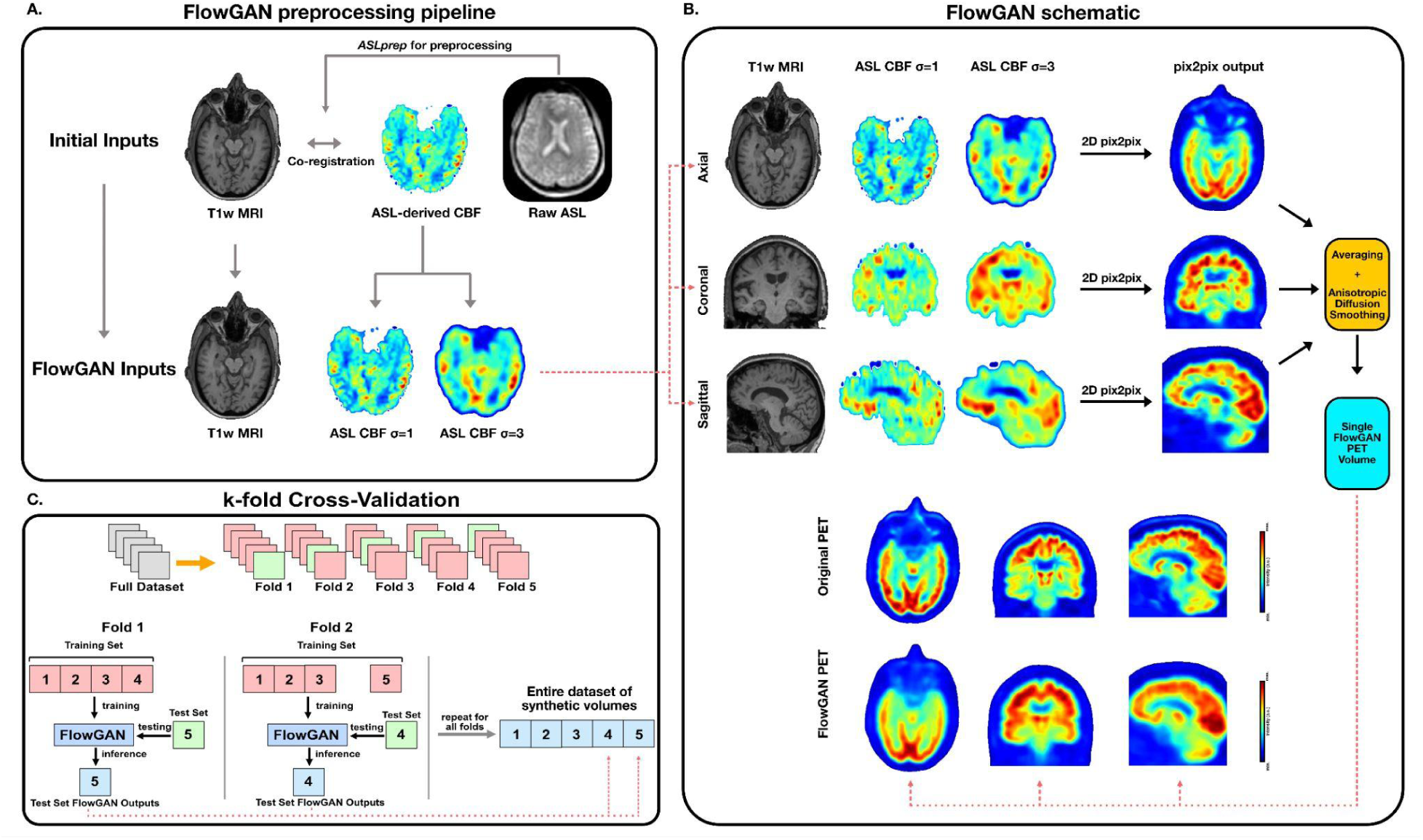
FlowGAN Pipeline. **Panel A.** shows the preprocessing pipeline that T1w and ASL inputs underwent before being fed into the FlowGAN pix2pix networks. The ASL data was preprocessed with ASLprep, which computed cerebral blood flow (CBF) maps and co-registered them to the T1w inputs. The resulting CBF maps were then duplicated and each was smoothed, one set with a Gaussian kernel of σ=1 and the other with a Gaussian kernel of σ=3, with each still co-registered to the T1w inputs. **Panel B.** illustrates the process of these inputs being fed into the three parallel pix2pix networks. The three input volumes were sliced into individual axial, coronal, and sagittal images, and the resulting 2D images were fed as a three-channel input into the FlowGAN pix2pix network corresponding to their plane (e.g., the axial images were fed into the pix2pix network trained on axial images) for inference. The resulting images were reconstructed into volumes for each of the three networks, yielding axial, coronal, and sagittal output volumes for each set of inputs. These output volumes were then themselves combined by averaging and smoothed using anisotropic diffusion to yield one final synthetic PET. **Panel C.** demonstrates the process of the k-fold cross-validation approach that was implemented when training and testing FlowGAN in order to both yield more robust results and increase the size of the test set while avoiding overfitting. This panel shows the process for 5 folds, but FlowGAN used a 12-fold cross-validation approach. The full dataset of 78 subjects was divided into 12 partitions, and in each of the 12 folds, 11 partitions were used for training and exactly 1 partition was left out as the test set and used for inference. All 12 partitions were used as the test set exactly once, yielding synthetic FlowGAN outputs for all 78 subjects.

### FlowGAN Training and Validation

#### Training

We developed LowGAN, a generative adversarial network (GAN) that enables low-field to high-field image translation, generating 3T-like brain MRI from T1w, T2w, and FLAIR inputs acquired on a 64mT scanner. Expanding this approach, we created FlowGAN to synthesize PET volumes from ASL and T1w MRI inputs. FlowGAN employs three parallel 2D pix2pix networks^25^, each handling different imaging planes (axial, coronal, and sagittal). Each network layer takes in three channels: T1w images and ASL-derived CBF maps smoothed with Gaussian kernels (σ=1 and σ=3) (**Figure 1B**). The resulting 2D outputs are stacked into a 3D volume. In LowGAN, a wavelet-Fourier filter was used to smooth the volumes from each plane^26^, which were then averaged together to produce the final outputs, but since PET images are inherently smoother, we opted for averaging and anisotropic diffusion smoothing.

To finalize the PET output, voxelwise averaging and anisotropic diffusion smoothing (kappa=80, gamma=0.1) were applied. Rigid registration corrected for minor shifts from model inference, aligning the synthetic image with the T1w space.

For downstream analysis, all volumes (T1w, ASL CBF, original PET, FlowGAN PET) were mapped to the 2009c asymmetric MNI template space. Using ANTsPyNet^23,24^, we skull-stripped each volume, resliced to the MNI template dimensions via Convert3D^27^, and registered using affine transformation with symmetric normalization.

FlowGAN was trained over 100 epochs with a batch size of 2 on 62 subjects (37 MCI, 25 CU) and validated on 16 left-out subjects (10 MCI, 6 CU), following LowGAN hyperparameter settings.

#### 12-fold Cross-validation

To address the limited size of the initial test set (16 subjects: 10 MCI, 6 CU), we implemented a 12-fold cross-validation scheme. This allowed us to train 12 separate models, each excluding 6–7 subjects from the training set (**Figure 1C**). Each subject was included in the training set of 11 models and served as the test subject for 1 model, which excluded them during training. This method enabled synthetic PET volume generation for all 78 subjects (47 MCI, 31 CU) while minimizing overfitting. Each pix2pix network across the 12 folds was trained for 100 epochs with a batch size of 2, following LowGAN hyperparameters.

#### Regional PET SUVR and ASL CBF values

All outputs, originally in the same native space, were normalized to the 2009c asymmetric MNI template space by first skullstripping, then reslicing, and then registering (using symmetric normalization with rigid, affine, and deformable transformations) the T1w MRI to the MNI template, and then applying the same transformations to the ASL CBF, FDG-PET, and FlowGAN PET images^28^. Utilizing three atlas segmentations—automated anatomical labeling (AAL)^29^, Desikan-Killiany-Tourville (DKT)^30,31^, and Harvard-Oxford (HO)^32,33^—standardized uptake values (SUV), cerebral blood flow (CBF), and regional voxel intensities were extracted from FDG-PET, ASL CBF, and T1-weighted images, respectively, for each region of interest (ROI) (**Figure 3A**). Voxel values were averaged for each ROI to compute absolute SUV, CBF, and intensity values. We then normalized these values to produce SUV ratios (SUVR), relative CBF values (rCBF), and relative voxel intensities, using the pons, putamen, or no normalization. In total, nine sets of SUVR, rCBF, and relative intensities were calculated across the three atlases and normalization methods.

### ROC-AUC Analyses Using MCI-CU Classifiers

#### All ROIs in Each Atlas

Each modality—T1-weighted (T1w) MRI, ASL CBF, original PET, and FlowGAN PET—was evaluated for its ability to classify participants as MCI or CU using a receiver operating characteristic (ROC) curve constructed via a random forest (RF) classifier with leave-one-out (LOO) cross-validation. For instance, a LOO RF classifier was trained on SUVR values derived from original PET images, using each region of interest (ROI), excluding CSF spaces, as a feature, with the area under the ROC curve (AUC) as a performance measure.

Since nine combinations of atlases (AAL, DKT, HO) and normalization methods (pons, putamen, none) were created, each was analyzed separately. For example, original PET SUVR, FlowGAN PET SUVR, ASL rCBF, and T1w relative voxel intensity normalized to the pons and using the AAL atlas were input to four separate LOO RF classifiers. The AUCs from each classifier were then compared to evaluate the ability of each modality to distinguish MCI from CU participants. This process was repeated across all atlas and normalization combinations (see **Supplementary Figure 1A**). Results are detailed in **Table 2**.

**Table 1.** Participant Demographics.

| **Demographics** | **Participant Demographics** |  |
| --- | --- | --- |
|  | **Group** |  |
|  | **Cognitively Unimpaired** | **MCI** |
| **Number of Subjects** | 31 | 47 |
| **Age in years** | 70.2 ± 6.9 (range: 56-83) | 73.0 ± 7.0 (range: 57-86) |
| **Sex (# female)** | 18 | 15 |
| **Education in years (median(range))** | 18 (11) | 17.5 (13) |
| **MMSE (median(range))** | 30 (5) | 27 (6) |

**Table 2.** Data from RF LOO classifiers using all ROIs. Results for each combination of atlas and normalization. **Atlas:** Automated Anatomical Labeling (AAL), Desikan-Killiany-Tourville (DKT), and Harvard-Oxford (HO). **Norm:** average values of ROIs for each modality were normalized to either the pons, putamen (put.), or not at all. **ROC AUC:** specific to the ROC curve generated by the classifier for the specific modality trained using the data for each feature in the atlas normalized to the normalization region. **95% CI:** 95% confidence interval for ROC AUC, as computed using DeLong’s test. **p-val:** Benjamini-Hochberg corrected p-value for a two-tailed test DeLong’s test assessing whether the ROC AUC for the modality is significantly different from the ROC AUC of the original PET. Bold indicates a p-value ≥ 0.05.

| Atlas | Norm | Original PET | | FlowGAN PET | | | ASL CBF ($\sigma=3$) | | | T1w | | |
| --- | --- | --- | --- | --- | --- | --- | --- | --- | --- | --- | --- | --- |
|  |  | ROC AUC | 95% CI | ROC AUC | 95% CI | p-val | ROC AUC | 95% CI | p-val | ROC AUC | 95% CI | p-val |
| AAL | Pons | 0.86 | 0.79, 0.95 | 0.58 | 0.46, 0.71 | $8.6 \times 10^{-5}$ | 0.63 | 0.51, 0.76 | $6.4 \times 10^{-4}$ | 0.80 | 0.71, 0.90 | **0.31** |
| | Put. | 0.79 | 0.71, 0.90 | 0.75 | 0.66, 0.87 | **0.66** | 0.54 | 0.40, 0.67 | $1.3 \times 10^{-3}$ | 0.77 | 0.69, 0.89 | **0.76** |
|  | None | 0.63 | 0.51, 0.76 | 0.81 | 0.71, 0.90 | **0.10** | 0.63 | 0.50, 0.75 | **0.93** | 0.64 | 0.50, 0.75 | **0.93** |
| DKT | Pons | 0.87 | 0.80, 0.95 | 0.71 | 0.59, 0.82 | 0.01 | 0.67 | 0.55, 0.79 | 0.01 | 0.80 | 0.71, 0.90 | **0.19** |
| | Put. | 0.84 | 0.77, 0.94 | 0.73 | 0.62, 0.84 | 0.02 | 0.58 | 0.45, 0.71 | $5.9 \times 10^{-4}$ | 0.79 | 0.70, 0.90 | **0.25** |
|  | None | 0.72 | 0.61, 0.83 | 0.82 | 0.73, 0.91 | **0.23** | 0.61 | 0.49, 0.74 | **0.23** | 0.66 | 0.54, 0.78 | **0.36** |
| HO | Pons | 0.85 | 0.76, 0.93 | 0.60 | 0.48, 0.73 | $1.4 \times 10^{-4}$ | 0.43 | 0.31, 0.57 | $7.3 \times 10^{-8}$ | 0.77 | 0.67, 0.87 | **0.18** |
| | Put. | 0.78 | 0.69, 0.89 | 0.71 | 0.60, 0.83 | **0.22** | 0.53 | 0.39, 0.65 | $3.2 \times 10^{-3}$ | 0.77 | 0.68, 0.88 | **0.93** |
|  | None | 0.68 | 0.57, 0.80 | 0.86 | 0.77, 0.94 | **0.06** | 0.63 | 0.49, 0.74 | **0.61** | 0.68 | 0.56, 0.79 | **0.89** |

To assess the efficacy of each modality as a biomarker, the highest ROC AUC value for each modality across combinations was noted (**Figure 3B**). For the four best classifiers, the top 10 most heavily weighted features were identified (**Figure 3D**). This analysis was conducted on both the FlowGAN test set (16 subjects) and the 12-fold cross-validation outputs on the full dataset (78 subjects).

#### Specific ROIs in Each Atlas

In addition to analyzing every region of interest (ROI) across the atlases, we also tested a set of seven hypothesis-driven features commonly predictive in FDG-PET studies: left and right hippocampi, left and right precunei, left and right posterior cingulate cortex (PCC), and a composite meta-ROI^12^. Using these regions, we repeated the classification analysis with LOO RF classifiers. ROC curves and AUC values are presented in **Supplementary Figure 1B**, with classifier results detailed in **Table 3**. **Figure 3C** shows the best ROC AUC for each modality across all combinations, while **Figure 3E** displays feature loadings for the top-performing classifier among the four modalities.

**Table 3.** Data from RF LOO classifiers using specific ROIs. Results for each combination of atlas and normalization when using only bilateral hippocampi, precunei, PCC, and meta-ROI as the features. **Atlas:** Automated Anatomical Labeling (AAL), Desikan-Killiany-Tourville (DKT), and Harvard-Oxford (HO). **Norm:** average values of ROIs for each modality were normalized to either the pons, putamen (put.), or not at all. **ROC AUC:** specific to the ROC curve generated by the classifier for the specific modality trained using the data for each feature in the atlas normalized to the normalization region. **95% CI:** 95% confidence interval for ROC AUC, as computed using DeLong’s test. **p-val:** Benjamini-Hochberg corrected p-value for a two-tailed test DeLong’s test assessing whether the ROC AUC for the modality is significantly different from the ROC AUC of the original PET. Bold indicates a p-value ≥ 0.05.

| Atlas | Norm | Original PET |  | FlowGAN PET |  |  | ASL CBF |  |  | T1w |  |  |
| --- | --- | --- | --- | --- | --- | --- | --- | --- | --- | --- | --- | --- |
|  |  | ROC AUC | 95% CI | ROC AUC | 95% CI | p-val | ROC AUC | 95% CI | p-val | ROC AUC | 95% CI | p-val |
| AAL | Pons | 0.84 | 0.77, 0.94 | 0.62 | 0.49, 0.74 | 3.3 x 10<sup>-3</sup> | 0.56 | 0.43, 0.69 | 5.0 x 10<sup>-5</sup> | 0.73 | 0.63, 0.85 | **0.05** |
|  | Put. | 0.76 | 0.67, 0.88 | 0.76 | 0.66, 0.87 | **0.88** | 0.60 | 0.46, 0.71 | **0.11** | 0.76 | 0.66, 0.87 | **0.88** |
|  | None | 0.72 | 0.61, 0.83 | 0.79 | 0.70, 0.90 | **0.28** | 0.56 | 0.43, 0.69 | **0.21** | 0.67 | 0.57, 0.80 | **0.67** |
| DKT | Pons | 0.84 | 0.77, 0.94 | 0.56 | 0.43, 0.69 | 1.2 x 10<sup>-4</sup> | 0.55 | 0.42, 0.68 | 2.7 x 10<sup>-6</sup> | 0.77 | 0.67, 0.87 | **0.09** |
|  | Put. | 0.78 | 0.69, 0.89 | 0.71 | 0.61, 0.83 | **0.56** | 0.54 | 0.40, 0.66 | 0.02 | 0.76 | 0.67, 0.87 | **0.80** |
|  | None | 0.63 | 0.51, 0.76 | 0.75 | 0.64, 0.86 | **0.44** | 0.64 | 0.53, 0.77 | **0.84** | 0.59 | 0.47, 0.72 | **0.84** |
| HO | Pons | 0.79 | 0.70, 0.89 | 0.64 | 0.52, 0.76 | **0.06** | 0.59 | 0.47, 0.73 | 7.8 x 10<sup>-3</sup> | 0.76 | 0.66, 0.86 | **0.55** |
|  | Put. | 0.72 | 0.63, 0.85 | 0.71 | 0.60, 0.83 | **0.79** | 0.54 | 0.41, 0.68 | **0.08** | 0.66 | 0.55, 0.79 | **0.56** |
|  | None | 0.66 | 0.55, 0.79 | 0.83 | 0.74, 0.92 | **0.11** | 0.65 | 0.53, 0.77 | **0.85** | 0.59 | 0.46, 0.71 | **0.41** |

This analysis was conducted on both the FlowGAN test set of 16 participants (10 MCI, 6 CU) and the 12-fold cross-validation outputs on the full 78-subject dataset (47 MCI, 31 CU).

### Statistical Analysis

We assessed original PET, T1w MRI, ASL CBF maps, and FlowGAN PET images using Structural Similarity Index (SSIM) and estimated correlations with Spearman’s rank correlation coefficients, setting statistical significance at p < 0.05. To manage multiple comparisons across brain regions, p-values were adjusted via the Benjamini-Hochberg False Discovery Rate (FDR) correction. ROC-AUC comparisons and confidence intervals were computed using DeLong’s test^34^. To evaluate batch effects on LOO RF classifier performance, we flattened PET, FlowGAN PET, ASL CBF, and T1w volumes into 1D vectors and applied t-Distributed Stochastic Neighbor Embedding (tSNE) (**Supplementary Figure 2**).

## Results

### FlowGAN visually recreates regional metabolism using T1w and ASL inputs

Demographic information for the study cohort is provided in Table 1. FlowGAN PET was derived from T1w MRI and ASL CBF inputs. In both MCI and CU subjects, FlowGAN PET volumes revealed hypometabolic patterns similar to true PET images, even though these differences are not visible in the original T1w and ASL inputs (Figure 1B, Figure 2). The structural similarity index (SSIM) between original PET and FlowGAN PET images was 0.958 (95% CI: [0.956, 0.960]). Red arrows in Figure 2 highlight hypometabolism in the hippocampi of MCI subjects compared to CU.

**Figure 2.**
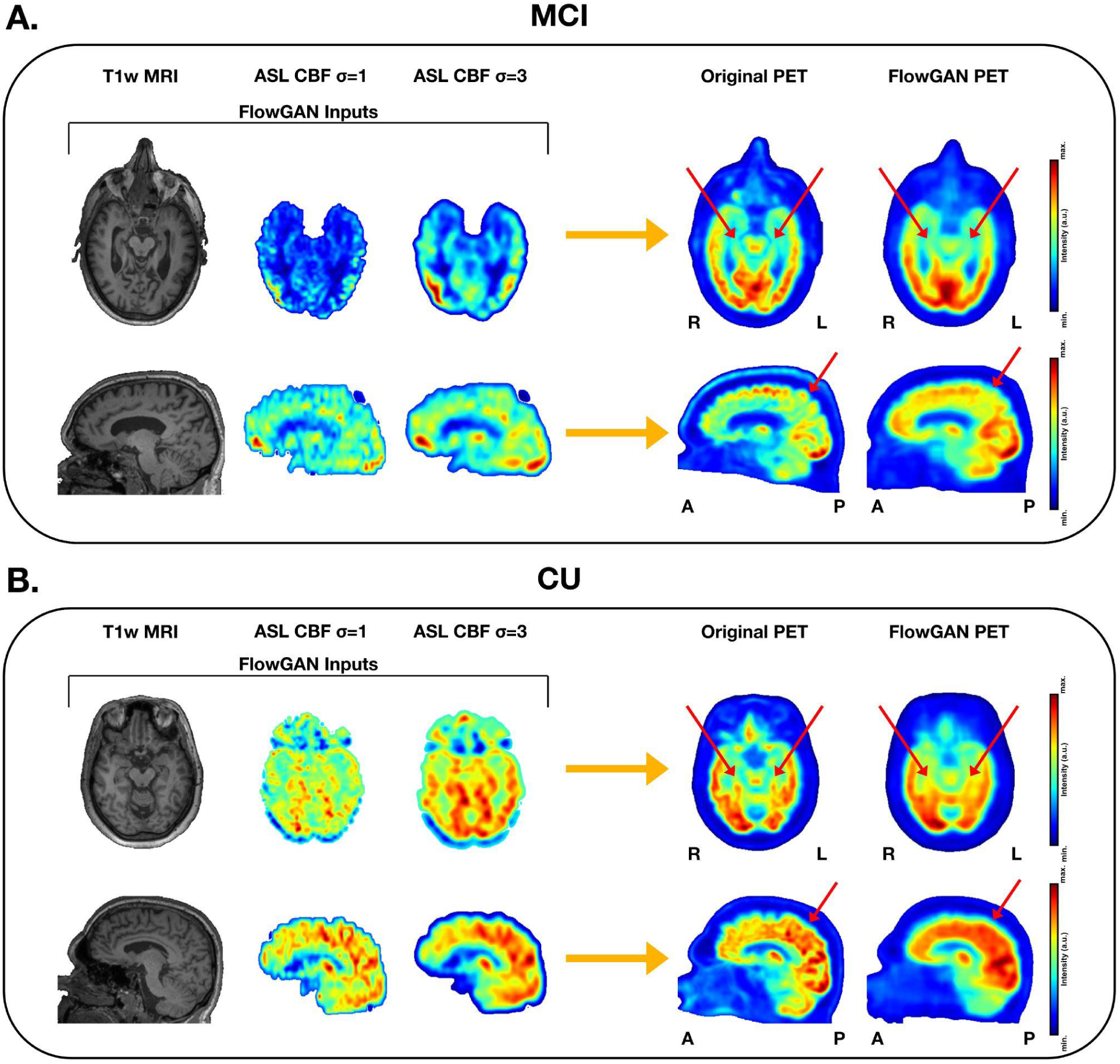
Representative FlowGAN outputs. On the left are the three FlowGAN inputs and on the right are the FDG-PET volumes, both original and synthetic. The red arrows point to the hippocampus and precuneus for the top and bottom subject, respectively. **Panels A. and B.** show MCI and CU subjects, respectively; MCI subjects demonstrate reduced bilateral hippocampal metabolism and precuneus metabolism, respectively, compared to CU subjects in their original PET volumes, which can also be seen in the synthetic volumes.

### FlowGAN PET performs comparably to true PET in classification

FlowGAN PET performed comparably to true FDG-PET in distinguishing MCI from CU participants when used to train a random forest classifier. Initially, all ROIs were used as classifier features. Under this configuration, the best classifier with original PET SUVR achieved an ROC-AUC of 0.87 (95% CI: [0.79, 0.95]) using the AAL atlas normalized to the pons (Figure 3B; Table 2). For FlowGAN PET, the top classifier had an ROC-AUC of 0.86 (95% CI: [0.77, 0.94]) using the HO atlas without normalization.

**Figure 3.**
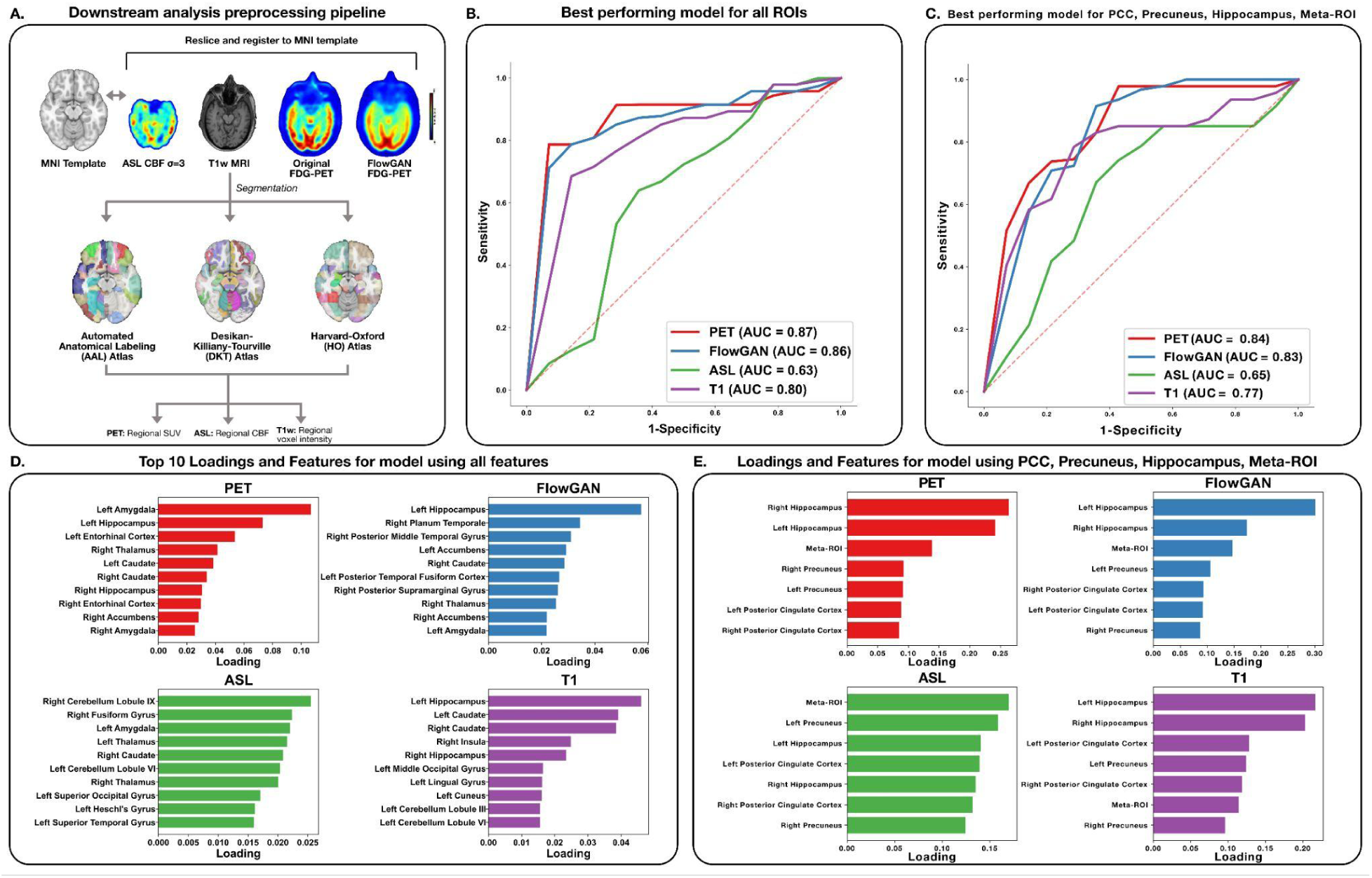
MCI vs. CU Classifier. **Panel A.** illustrates the preprocessing pipeline to prepare the data for the classifier. The T1w, ASL CBF (σ=3), original PET, and FlowGAN PET were mapped to the 2009c asymmetric MNI template space. Three separate segmentations of this space—AAL, DKT, and HO atlases—were applied to each volume to segment the volumes into ROIs. The data were then duplicated, and for one set, the average values—SUV for PET, CBF for ASL, voxel intensity for T1w—over every ROI in the atlas were used as the features for the classifier, for the other set, only bilateral hippocampi, precunei, PCC, and meta-ROI were included. These features were also normalized in three different ways—to the pons, to the putamen, or not at all—which generated nine total datasets for each modality in both the cases of using all ROIs and only the specific ROIs. The RF classifiers were trained in a LOO method and were then used to construct an ROC curve for each modality. **Panel B.** shows the best ROC curve (greatest AUC) for each modality when all ROIs (excluding CSF spaces) in each atlas were included as the features. **Panel C.** exhibits the best ROC curve for each modality when only the aforementioned specific ROIs were included as features. In addition to this analysis, the loadings (importances) of each classifier were investigated, which showed the contribution of each feature to the classifier’s prediction. **Panel D.** presents the 10 features with the greatest loadings and said loadings for the best classifier for each modality when all ROIs were included as features. **Panel E.** produces all of the features and their loadings for the best classifier for each modality when only specific ROIs were included as the features.

ASL CBF (σ=3) reached an ROC-AUC of 0.67 (95% CI: [0.55, 0.79]) with the DKT atlas normalized to the pons, and T1w MRI achieved 0.80 (95% CI: [0.71, 0.90]) using the AAL atlas with pons normalization. We additionally examined the top 10 loadings for the features of each of the best performing models to investigate their relative contributions to the models (**Figure 3D**).

Given that canonical ROIs for differentiating MCI or AD from CU based on either FDG-PET or ASL MRI include bilateral hippocampi, precunei, PCC, and a meta-ROI^12^, we also trained classifiers using only these 7 regions. The best original PET SUVR classifier using these regions achieved an ROC-AUC of 0.84 (95% CI: [0.77, 0.94]) with the AAL atlas normalized to the pons. Similarly, the top FlowGAN PET classifier reached an ROC-AUC of 0.83 (95% CI: [0.74, 0.92]) using the HO atlas without normalization, while ASL CBF and T1w MRI classifiers achieved ROC-AUCs of 0.65 and 0.77, respectively (**Figure 3C**). Feature loadings for the best classifiers are detailed in **Figure 3E**.

Since T1w MRI classifiers performed nearly as well as FlowGAN PET and better than ASL CBF, we hypothesized that T1w MRI inputs contributed more to FlowGAN than ASL CBF. We thus trained a FlowGAN variant with only T1w inputs, excluding ASL CBF. Using all ROIs, the best classifier for this new model achieved an ROC-AUC of 0.82 (95% CI: [0.74, 0.92]) with the DKT atlas, while the focused ROIs classifier reached 0.81 (95% CI: [0.72, 0.90]).

Finally, tSNE analysis was conducted on original T1w MRI, ASL CBF (σ=1 and σ=3), original PET, and FlowGAN PET to assess batch effects, revealing no significant clustering (**Supplementary Figure 2**).

## Discussion

The findings of our study demonstrate that synthetic FDG-PET derived from T1w MRI and ASL MRI (FlowGAN PET) generates not only visually similar metabolic patterns observed in true PET images of MCI participants (often perceptually absent in T1w MRI and ASL CBF), but also performs comparably to true FDG-PET in distinguishing MCI from CU subjects from using regional SUVRs. Notably, the most influential features of the classifiers were also consistent between true PET and synthetic PET. Our results also demonstrated that T1w MRI classifiers alone outperformed ASL CBF-based classifiers and nearly matched FlowGAN and true PET accuracy, likely reflecting the poor quality of 2D ASL MRI data used in this study. A modified FlowGAN using only T1w MRI slightly underperformed compared to FlowGAN using both T1w and ASL CBF inputs. Interestingly, feature loadings differed between T1w-only FlowGAN outputs and those using both inputs, suggesting that while T1w MRI provides structural data, ASL CBF adds unique perfusion information that enhances synthetic PET quality.

To ensure these findings were not an artifact of any one parcellation, we repeated the entire analysis with three complementary brain atlases (AAL, DKT, and HO). Using multiple atlases reduces parcellation bias, offers broader anatomical coverage, and tests the network across ROI sizes that differ in their sensitivity to partial-volume and registration error^35^. The overall pattern of FlowGAN PET being comparable to true PET, both of which were better than T1w MRI, and all three of which were better than ASL CBF, held across atlases, although individual AUC values varied as expected from differences in ROI granularity and reference-region definitions.

While the best-performing classifiers using true PET were normalized to the pons (consistent with literature), those for FlowGAN required no normalization. This may be due to the input normalization across the volume before inference as part of the FlowGAN architecture, making post-normalization redundant and potentially limiting.

Additionally, tSNE analyses failed to demonstrate batch effects, given no clustering by condition. These findings reinforce the ability of FlowGAN to capture metabolic patterns from T1w and ASL inputs, translating these into PET-like images that can effectively differentiate MCI from CU participants. Our study highlights the robustness of FlowGAN and LowGAN frameworks and suggests broader potential applications.

Limitations remain, however. Although FlowGAN-generated PET volumes performed comparably to real PET in differentiating MCI from CU at a group level, these synthetic images have yet to undergo evaluation at the single subject level by neurologists and neuroradiologists.

### Conclusion

This study extends the utility of FlowGAN from diagnosis and monitoring of TLE to the AD-continuum. Given their similarity and comparable performance of synthetic FlowGAN PET to true FDG-PET in differentiating MCI from cognitively unimpaired subjects, synthetic FDG-PET derived from T1w MRI and ASL MRI shows promise in improving the utility of ASL MRI in the differential diagnosis of dementia.

## Data Availability

Upon publication of our manuscript, we will release our GitHub repository with code to train FlowGAN, as well as the pre-trained weights used in this manuscript. We will also release the code used to generate the figures and the analysis described in this manuscript.

