## Supplemental Figures for "Improving the Detection of Mild Cognitive Impairment with FlowGAN: a Framework for ASL to FDG-PET Image Synthesis"

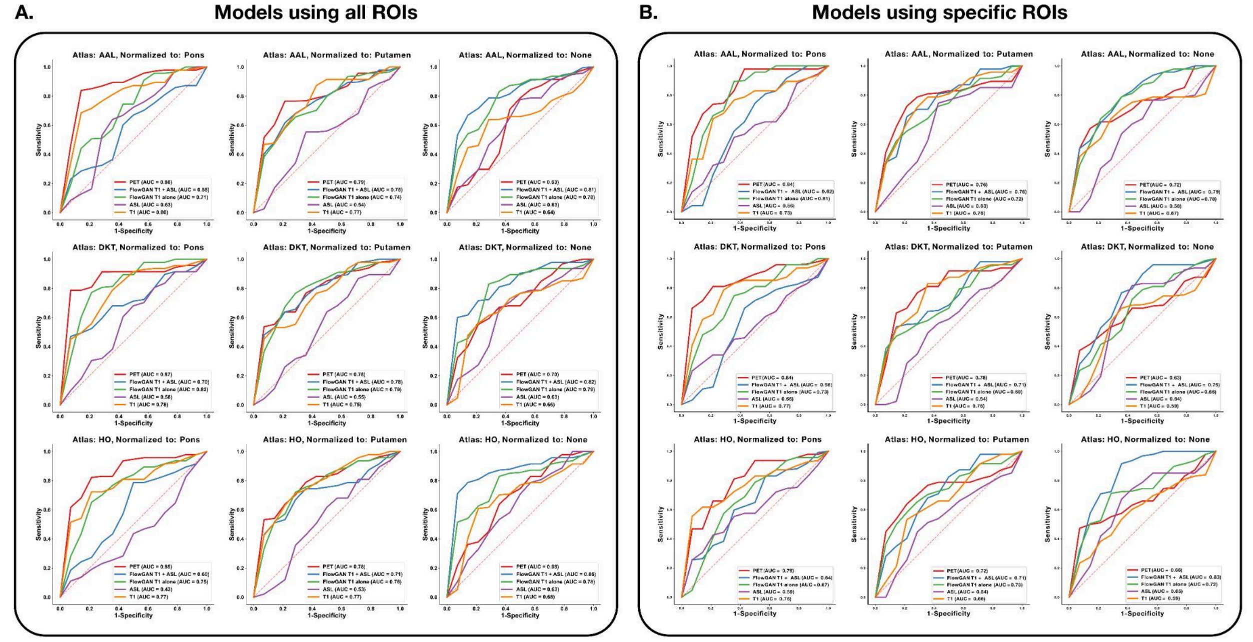


**Supplementary Figure 1 — All classifiers created from 12-fold cross-validation outputs:** Given that 3 atlases were used—AAL, DKT, and HO—and the 3 types of normalization—pons, putamen, and none at all—there were a total of 9 combinations of atlases and normalizations. For each combination of atlas and normalization, 5 RF classifiers with LOO cross-validation were created using the data from a single modality: original PET, synthetic FlowGAN PET trained using both T1w MRI and ASL CBF, synthetic FlowGAN PET trained using T1w MRI alone, ASL CBF (σ=3), and the original T1w MRI. **Panel A.** shows the ROC curves and their respective AUC values when every ROI in the atlas was given as a feature for the RF classifier. **Panel B.** depicts the ROC curves and their respective AUC values when only the bilateral hippocampi, precunei, PCC, and meta-ROI were provided as features for the classifier.

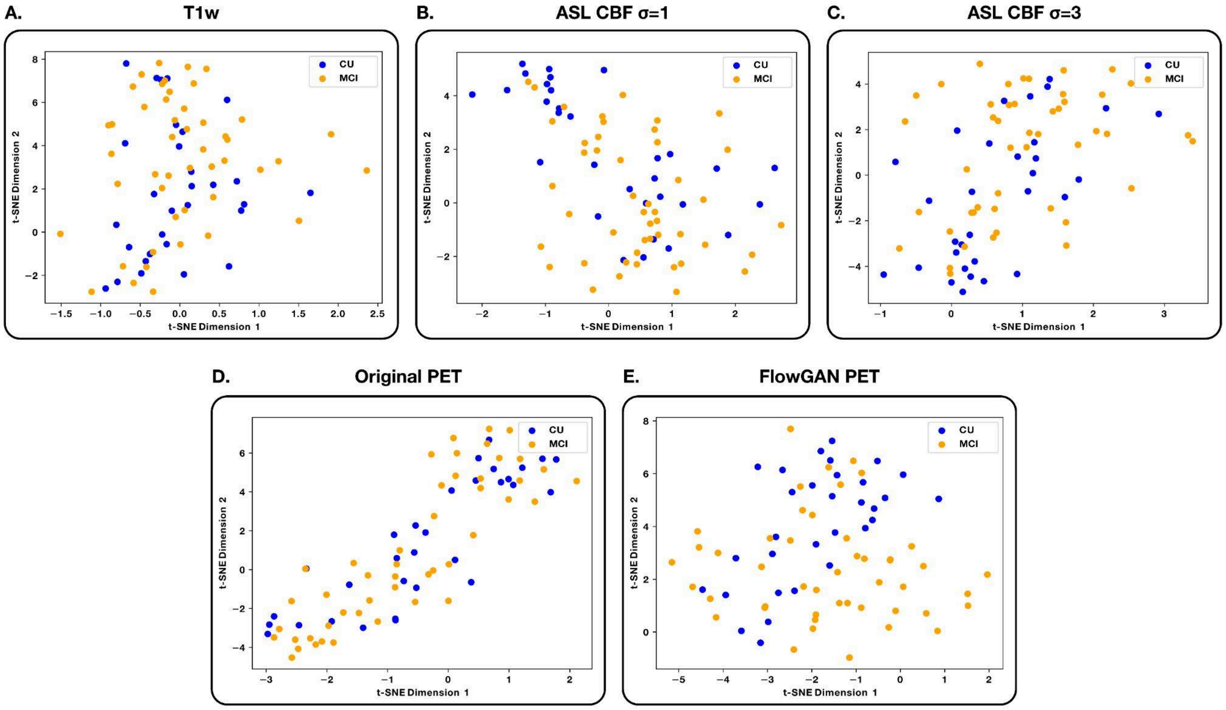


**Supplementary Figure 2 — tSNE plots for each modality:** For each input modality—T1w MRI, ASL CBF (σ=1), and ASL CBF (σ=3)—as well as the PET volumes—original and FlowGAN—tSNE plots were created in order to test for potential batch effects within the data. **Panels A-E** show the tSNE plots for the T1w MRI, ASL CBF (σ=1), ASL CBF (σ=3), original PET, and FlowGAN PET, respectively.
